# Efficient Synthesis of 3D MR Images for Schizophrenia Diagnosis Classification with Generative Adversarial Networks

**DOI:** 10.1101/2024.06.01.24308319

**Authors:** Sebastian King, Yasmin Hollenbenders, Alexandra Reichenbach

## Abstract

Schizophrenia and other psychiatric disorders can greatly benefit from objective decision support in diagnosis and therapy. Machine learning approaches based on neuroimaging, e.g. magnetic resonance imaging (MRI), have the potential to serve this purpose. However, the medical data sets these algorithms can be trained on are often rather small, leading to overfit, and the resulting models can therewith not be transferred into a clinical setting. The generation of synthetic images from real data is a promising approach to overcome this shortcoming. Due to the small data set size and the size and complexity of medical images, i.e. their three-dimensional nature, those algorithms are challenged on several levels. We develop four generative adversarial network (GAN) architectures that tackle these challenges and evaluate them systematically with a data set of 193 MR images of schizophrenia patients and healthy controls. The best architecture, a GAN with spectral normalization regulation and an additional encoder (α-SN-GAN), is then extended with an auxiliary classifier into an ensemble of networks capable of generating distinct image sets for the two diagnostic categories. The synthetic images increase the accuracy of a diagnostic classifier from a baseline accuracy of around 61% to 79%. This novel end-to-end pipeline for schizophrenia diagnosis demonstrates a data and memory efficient approach to support clinical decision-making that can also be transferred to support other psychiatric disorders.

## Introduction

Schizophrenia (SCZ) is a heterogeneous neurological disease characterized by a broad spectrum of symptoms including delusions, hallucinations, and disorganized thinking [1]. Due to a lack of reliable diagnostic biomarkers [1, 2], psychiatrists currently diagnose SCZ based on the Diagnostic and Statistical Manual of Mental Disorders (DSM-V) [3]. The disorder is connected to a variety of genetic and environmental factors and genetic, blood, as well as brain alterations have been linked to it [1, 4].

Brain imaging with structural magnetic resonance imaging (sMRI) has been explored for supporting the objective diagnosis of SCZ. Due to the widespread but subtle changes in brain matter of SCZ patients [5], multivariate approaches such as machine learning (ML) algorithms are prominently explored for automated support of objective SCZ diagnoses. ML considers a diagnosis as classification problem sorting images into the classes „healthy“ or „patient“. Approaches include feature extraction combined with traditional ML algorithms [5] as well as deep learning (DL) algorithms, which allow automatic feature extraction [6, 7]. The latter enables SCZ classification without a priori hypotheses about specific brain regions being discriminatory for the task at hand. 3D convolutional neural networks (3D-CNNs), a type of DL algorithms for three-dimensional inputs, achieve state-of-the-art performance up to 95% accuracy for SCZ “diagnosis” based on sMRI images [7-9]. However, DL algorithms tend to overfit on small data sets [10].

Training robust and reliable DL classifiers requires large training data sets, which is challenging for medical image data. Image acquisition is expensive and expert knowledge is required to label the data. Therefore, an effort is made to assemble such images in publicly available data sets to further research. However, publicly available MRI data sets from SCZ patients and healthy controls (HC) are still rather small, ranging from 50 to 600 images per class [8]. To increase the robustness of classification, data augmentation techniques have been employed to bolster small data sets for training the classifier [11]. Traditional approaches from image classification, e.g. affine transformations or similar distortions, are less suitable for MRI images. However, generative ML methods like generative adversarial networks (GAN) provide promising new techniques for data augmentation [12]. An added bonus of these techniques is the generation of genuinely synthetic data, which does not underlie the strict regulations of patient data [12, 13]. The data generated with these models can therefore be published more easily and benefit a wide crowd of researchers. GAN architectures consist of at least two neural networks: a generator and a discriminator. The generator tries to produce images similar to those of a reference data set by using the feedback from the discriminator who learns to distinguish between real images and synthetic ones. GANs in various adaptions have shown the technical capability of synthesizing medical imaging data including the generation of brain images [13], mainly for 2D slices but also for 3D volumes [12, 14, 15]. They can also be conditioned to produce data of a given class affiliation [12, 16, 17], i.e. patients or HC. Medical images like sMRI volumes challenge GANs four-fold: 1) small data sets can lead to discriminator overfit, causing the vanishing gradients problem [18, 19]; 2) most algorithms for image processing were first, or only, implemented and optimized for 2D data; 3) complex, i.e. 3D, images tend to converge to a very small distribution of generated images causing mode collapse [14]; 4) 3D images and operations need exponentially more memory. End-to-end generation of synthetic data from a very small 3D sMRI data set and downstream data-driven “diagnosis” classification has not yet been demonstrated, nor have MR images for SCZ classification been produced before.

We address the previously mentioned challenges with a publicly available data set of sMRI data consisting of 102 SCZ patients and 91 HC, furthering the development of four 3D deep convolutional GAN (DC-GAN) architectures with various modifications, and systematically comparing the resulting architectures. The winning architecture is then extended by three conditional approaches to produce data from the two clinical groups and the best combined architecture is chosen to produce synthetic data for training a 3D-CNN SCZ “diagnosis classifier”. This classifier is trained with different ratios of real and synthetic data and then tested with real data. An increase in classification accuracy when training with synthetic data demonstrates the added value of this generated data. In sum, this study contributes to the data and memory-efficient training of DL classifiers in medical imaging.

## Methods

### Data

The data set used in this study was obtained from the MCIC collection [20] in July 2019. The collection contains structural T1-weighted MR images of 158 adult SCZ patients and 169 demographic, age, and sex-matched HC. Four research sites were involved in the data collection process from 2004 to 2006. All subjects provided informed consent to participate in the study that was approved by the human research committees at each of the sites. Patients had to be diagnosed with SCZ conforming to the Diagnostic and Statistical Manual of Mental Disorders, 4th Edition (DSM-IV). A differentiation between distinct types and severities of SCZ was not conducted. We included only data from sites A, C, and D because the images originating from site B were not publically released due to IRB restrictions. Furthermore, the data from nine subjects failed transformation to BIDS format [21] due to missing meta-data, leaving a subset of 102 SCZ and 91 HC for this study. Sex distribution in the remaining data set was imbalanced with 61 female (A: 20; C: 21; D: 20) and 152 male (A: 70; C: 39; D: 43) subjects. Age of the subjects ranged from 18 to 60 years (A: 18-60; C: 18-60; D: 20-57).

Data were pre-processed using the nypipe toolbox [22] for Python. First, images were skull-stripped and registered to MNI space in 1mm^3^ isotropic resolution with nypipe wrapper functions for SPM12 [23]. To remove outliers, we capped voxel intensity at upper and lower 1% quantile of values and then rescaled the images to the range of -1 to 1. Finally, slices containing only background were trimmed.

In order to reduce the complexity of the classification and generation problem, we tested in a pre-study whether the reduction of image size from 128^3^ to 64^3^ voxel compromised the information content usable for a diagnostic classifier. Using the same classification algorithm and training strategy as in the main study (cf. Diagnosis classifier), we found that classification accuracy did not suffer from down sampling (59.1±7.1% for high resolution and 60.6±7.6% for low resolution; *t_8_*=0.29; *p*=.779). The number of parameters, however, decreased around 12-fold from 90,198,561 to 7,361,057, analogously decreasing the computational resources for training the classifier. Therefore, we proceeded with images sampled down to around 2mm^3^ voxel size with 64^3^ voxel for the main study.

### Experimental procedure

Four GAN architectures based on a 3D DC-GAN are adapted to address our challenges, and evaluated for their image synthesis capabilities (Figure 1). Spectral normalization regularization (SN-GAN) [24] counteracts the vanishing gradients problem that often occurs for small sample sizes and is therefore applied for all architectures. Additionally incorporating an encoder (α-SN-GAN) [14] helps to alleviate mode collapse.

**Figure 1.**
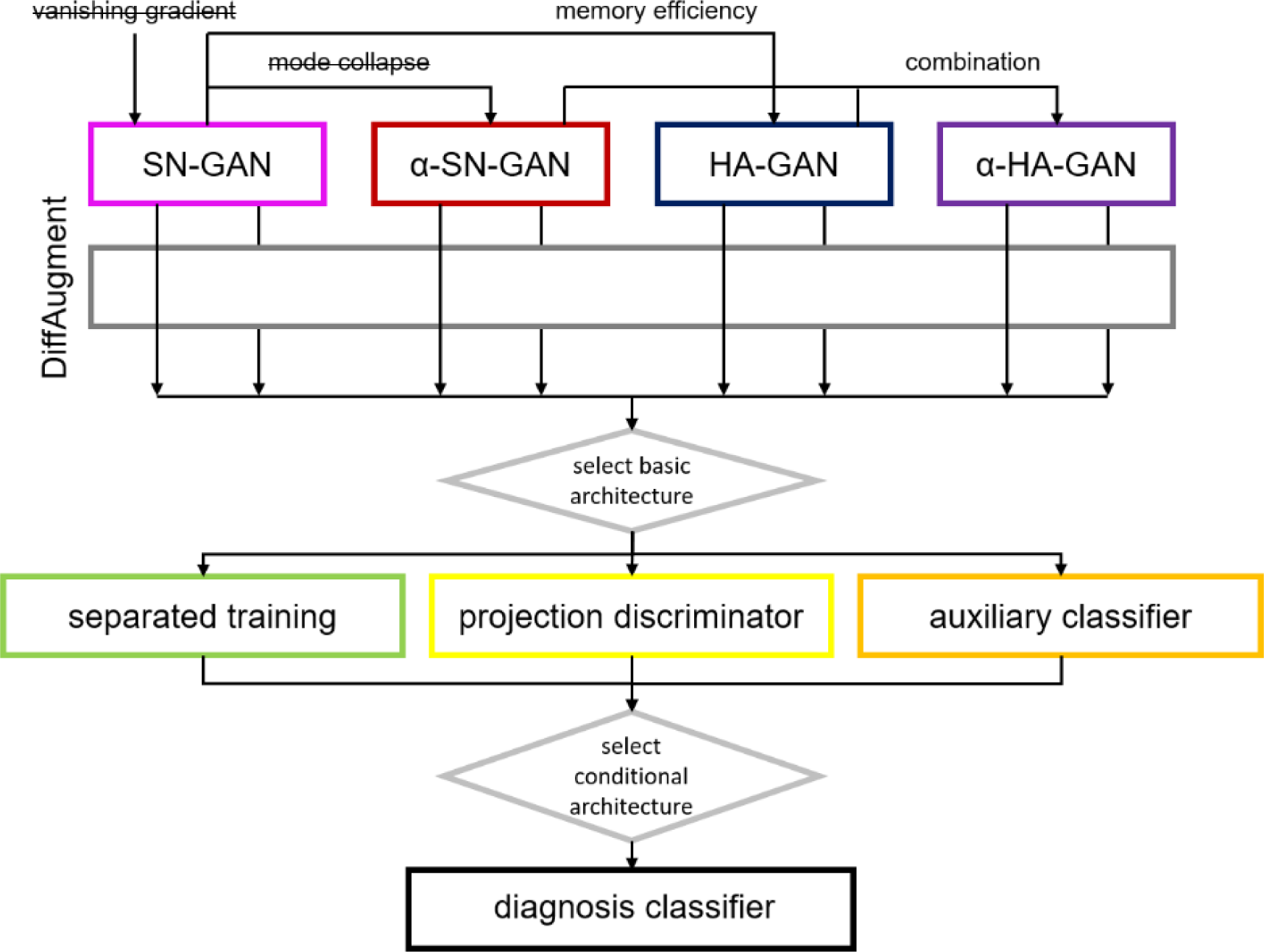
Processing pipeline with architecture selection. Four basic architectures are designed to alleviate different problems during data synthesis and all four are tested with and without additional augmentation during training (DiffAugment). The best basic architecture is then expanded by three approaches for the generation of different clinical classes (schizophrenia patients *vs.* healthy controls). The best conditional architecture is then used to synthesize data for training a classifier that can be used for decision support in schizophrenia diagnosis. Abbreviations: SN (spectral normalization), GAN (generative adversarial network), α (with encoder), HA (hierarchical amortized)

To reduce the computational cost of the training, a hierarchical approach is adapted (HA-GAN) [15], which is also combined with the α-SN-GAN to join their advantages (α-HA-GAN). Vanishing gradient and mode collapse are additionally addressed by applying data augmentation during training (*DiffAugment*) [19] to all four architectures, resulting in eight basic architectures being tested. The best architecture is selected for further processing based on qualitative and quantitative evaluation.

Subsequently, three conditioning approaches are employed for creating images of the two clinical groups (SZC / HC): one classifier per class, an auxiliary classifier, and a projection discriminator. The winner architecture is then used to generate different sets of training data with different ratios of real and synthetic data and different set sizes. Finally, a diagnosis classifier is trained on these data sets to separate SCZ patients from HC in a test data set consisting of real data only.

### Diagnosis classifier

The purpose of generating synthetic data in this study is to train a classifier that separates sMRI from SCZ and HC, henceforth called a diagnosis classifier. We employed a 3D-CNN since these types of networks have shown high performance in diagnosing schizophrenia from sMRI in previous studies [7-9, 25]. The architecture is based on the VGG16 architecture [26]. In brief, the model consists of multiple 3D convolution blocks and linear layers at the end. A 3D convolution block consists of a convolution layer, a Rectified Linear Unit (ReLU) activation function, a batch normalization layer, and a max pooling layer. In between the linear layers, dropout is used. For details on the architecture, see Supplementary material Figure S1.

For the training of each classifier, we chose a stratified 5-fold cross-validation approach with a batch size of 12. The training is executed with an AdamW optimizer [27] with a learning rate of 5e^-6^ and weight decay of 0.01. Hyperparameters are based on [26] and then tuned manually. To account for overfitting effects, all classifiers are trained past their convergence and the classifier at the minimum validation loss of each split is chosen.

### Basic architectures for image synthetization

GAN architectures in this study are all based on the standard deep convolutional GAN (DC-GAN) [28] architecture, sometimes also referred to as CNN-GAN. This architecture consists of two networks. The generator network synthesizes images that resemble real sMRI images by projecting a randomly sampled latent space of a prior distribution z to the target distribution X. The discriminator network learns to distinguish real sMRI images from synthetic ones, produced by the generator. The adversarial loss consists of two different loss terms for updating the discriminator and the generator. Despite its success, the DC-GAN architecture suffers from two major problems: vanishing gradients [29, 30] and mode collapse [14, 24, 29, 30]. Vanishing gradients describes a phenomenon in which the discriminator network saturates quickly and in return fails to provide a meaningful gradient to the generator. This problem is magnified for small data sets since they are easily memorable for the discriminator. Mode collapse occurs when the generator succeeds in creating images that are classified as real by the discriminator but only produces a small variety of them.

All GANs were trained using the Adam optimizer with a learning rate of 0.0002, β1=0.5, and β2=0.999 for all networks. We chose those training parameters and a latent space size of 1000 since this size has shown to be effective in related works [14, 15]. Each GAN training was performed for 12,000 iterations with a batch size of four.

#### SN-GAN

A DC-GAN [23] with Spectral Normalization (SN) [24] applied (SN-GAN) is used to alleviate the vanishing gradients problem. Two changes regarding the discriminator regularization and the upsampling in the generator are applied to the DC-GAN architecture to form the SN-GAN (cf. Supplementary material Figures S2 and S3). The basic idea of regularization techniques is to avoid steep gradients and therefore fast changes in the discriminator’s weights, which lead to the vanishing gradients problem. Spectral normalization achieves this by renormalizing the weights of a layer according to 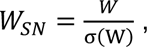 where σ denotes the spectral norm of the weight matrix *W*. This enforces a maximum Lipschitz constant of 1 for the discriminator which results in slower but more stable updates of the network.

SN has achieved competitive results over the application of a gradient penalty (GP) [29], which is the successor to weight clipping in Wasserstein GANs (WGAN) [30]. Contrary to SN, GP only penalizes large gradients by adding a penalty term to the loss function instead of enforcing a hard constraint. Due to its lower computational cost compared to GP, SN is preferred over GP in this study for all models regardless of their original regularization technique.

Another change compared to the original DC-GAN addresses the upsampling technique in the generator. Instead of transposed convolutions, we used pixel-shuffle [31] layers to increase the image size. This technique is chosen because it is computationally more efficient while increasing the number of parameters by shifting the upsampling process to the feature dimension. Contrary to interpolation approaches for upsampling, pixel-shuffle allows for learned upsampling filters and therefore leads to a higher flexibility of the network. To eliminate the occurrence of checkerboard artefacts, we use an initialization scheme that resembles nearest-neighbour upsampling in the early stages of the training [32]. To further increase the flexibility of the model, the leaky ReLU activation function is used in the generator as well as the discriminator. The original DC-GAN implementation uses the ReLU activation function only in the generator.

#### α-SN-GAN

To combat mode collapse, an α-WGAN-GP [14] architecture, which has been used for medical 3D image generation on small medical data sets, is employed. We adapt the architecture of the α-WGAN-GP with SN and replace the Wasserstein Loss [30] with the Hinge Loss [24, 33], therefore naming the adapted architecture α-SN-GAN.

To encourage the generator not to fall into the mode collapse, its goal is changed by introducing an encoder to the architecture [14, 34, 35] (cf. Supplementary material Figure S4). The encoder is used to produce a latent space representation of real images, which is used for training the generator along with the randomly sampled latent spaces. An additional reconstruction loss term like the L1 loss encourages the generator to produce images that cover the diversity of the whole data set [34, 35]. The loss function of the encoder is based on the Kullback-Leibler divergence [35] as distance metric, which leads the encoder to produce a distribution close to a randomly sampled latent space.

To counteract a common drawback of GAN architectures with encoders, blurry images, we adapted the α-GAN approach [34] for all encoder models in this study. Our approach introduces a code discriminator (cf. Supplementary material Figure S5) discriminating between randomly sampled latent spaces and encoded latent spaces. The code discriminator considers the randomly sampled latent spaces as real and the encoded latent spaces as fake, therewith encouraging the encoder to generate latent spaces close to the random distribution. The code discriminator used in this study consists of linear layers with spectral normalization. It is trained with the same loss function as the regular discriminator.

#### HA-GAN

To achieve resource efficiency, we leveraged the hierarchical HA-GAN [15], which was proposed to synthesize 3D medical images computationally effectively. It achieves shorter training times at a reduced memory consumption compared to the architectures previously introduced.

The HA-GAN implements the hierarchical structure by splitting each of the GAN’s networks into a high-resolution and a low-resolution path (cf. Supplementary material Figure S6). A random image sub-volume of fixed size is used to train the high-resolution path whereas the low-resolution path is trained using a down-sampled version of the image thus lowering the overall memory consumption. In addition to these models, both generators share a generator that synthesizes images up to the size of the low-resolution images, whose output is then used for the high- and low-resolution generators. During inference mode, when no gradients need to be calculated, the high-resolution generator synthesizes full-sized images.

We adapted the HA-GAN architecture to create 64³ images with a sub-volume size of 8x64x64 and down-sampled images to one fourth of the original size (16³). The HA-GAN has been introduced for larger data sets and larger image sizes (128³ and 256³) [15], on which it demonstrated improved quality compared to a WGAN and α-GAN. The models’ performance on a small data set as well as smaller image size has not yet been tested.

#### α-HA-GAN

The combination of the α-SN-GAN’s training scheme and the HA-GAN architecture offers the possibility of performing a resource-efficient training of 3D GANs on small data sets (α-HA-GAN). The α-SN-GAN’s encoder is replaced with the memory-efficient encoder [15] and in the same way the discriminator and generator are split into a high- and low-resolution network. The high-resolution encoder produces a latent space of the same size as the high-resolution generator’s input and is trained by an L1 loss to minimize the difference between the input image and the high-resolution generator’s reconstruction of it. A concatenation of all high-resolution encoder outputs of all non-overlapping sub-volumes X_c_ of the image is fed into the low-resolution encoder, which then is trained in the same fashion as the α-SN-GAN’s encoder. The resulting network combines all previously introduced techniques to leverage their improvements (Supplementary material Figure S6).

Contrary to the SN-GAN and the α-SN-GAN architectures, the hierarchical ones are trained using the standard GAN loss since the Hinge Loss led to instability in the training.

#### DiffAugment

Additionally to the four architectures, we adapted DiffAugment [19] for 3D images and applied this technique to each architecture. DiffAugment describes the application of data augmentation in the training process of the GAN itself boosting the data set with augmented images to counteract mode collapse as well as the vanishing gradients problem. Augmentations are achieved with translation, masking, and colour or intensity range bias in the images. These augmentations are applied to each input of the discriminator. Since the generator is updated using the discriminator’s feedback, its gradients need to be able to flow through the discriminator as well as the augmentations during the backward pass of the generator. It is therefore necessary for the applied augmentations to be differentiable. Differentiable augmentations are applicable for real and synthesized discriminator inputs contrary to regular augmentations. This is necessary to prevent augmentation artefacts in the synthesized images.

### Architectures for conditional image synthetization

The purpose of data synthetization in this study is to generate training data for diagnosis classification; therefore, we need to generate data for two classes, i.e. two clinical groups: SCZ and HC. The most straightforward approach is to train two separate GANs with data from each group, respectively. A complementary approach is employed with conditional GANs (cGANs) [16] incorporating a class label in the synthetisation process. Different approaches exist for introducing this information in the generator and the discriminator. For the generator, we introduce the class information by concatenating an embedding of the label to the latent space for all conditional models. For the discriminator, we implement two different approaches.

#### Projection discriminator

Conditional discriminators incorporate the label in their architecture and output only the realism score or probability of being real for a given input [16, 36-38]. Three techniques have been proposed to introduce the class label y to the discriminator: Concatenation of the label to the input layer [16], to an intermediate layer [37, 38], or alternatively forming the inner product to include the label information instead of the concatenation, resulting in the projection discriminator [36]. The latter has previously demonstrated the highest performance [36] and will therefore be used in this study.

#### Auxiliary classifier

Auxiliary Classifier GANs (AC-GANs) [39, 40] use an additional classification to provide feedback about the distinctness of the two data sets to the generator. They can be approached in two ways. The first one splits the discriminator at an intermediate layer to perform two different classifications: one for the realism of the image and one for the class affiliation [39]. The second approach involves a separate classifier [40]. In the latter, the original discriminator remains unaltered, thus its architecture is similar to a non-conditional discriminator. In theory, this method yields better results since the separated classifier is only trained on real images and is therefore not influenced by generated images. Additionally, unbalanced data sets are accounted for by the separated classifier contrary to the first approach [40]. These reasons lead to the use of only the second approach in this study. The auxiliary classifier is a 3D-CNN classifier using the same structure as the discriminator (cf. Supplementary material Figure S3). It was trained in advance to overfit on the real data set to ensure correct classification of synthetic images. During the GAN training it is only used in inference mode and not trained any further.

### Evaluation of synthetic data

#### Quantitative evaluation

Since the goal of GAN generators is the synthesize data with a distribution similar to the real data distribution, we utilize distance metrics to quantify the synthetization quality. Two commonly used distance metrics are the Maximum Mean Discrepancy (MMD) [41] and the Fréchet Inception Distance (FID) [41]. Both metrics combine the fidelity and diversity of generated images into one single score. To separate those performance measures, precision and recall metrics [42] are additionally calculated. Precision represents the proportion of generated images inside the target distribution whereas recall represents the coverage of the target distribution by the generated images.

In most image generation studies, GAN distance metrics are calculated based on features extracted with an Inception network architecture [43] pre-trained on the ImageNet data set [44] rather than whole images [41, 42, 45]. For medical 3D MR images, however, we could not use this pre-trained model because the Inception network cannot process 3D image data and a classifier trained on the ImageNet data set does not extract medically relevant features [46]. To alleviate both problems, a ResNet50 architecture pre-trained on 3D medical image data sets [47], was adapted to perform the extraction that were then used to calculate all the distance metrics. Note that this approach prevents the direct comparability of this study to others that use those distance metrics based on the Inception network trained on the ImageNet data set.

For each of the trained architectures, we generate ten data sets each containing as many synthetic images as real images available for the respective generation model.

#### Qualitative evaluation

The synthetic data is evaluated qualitatively based on exemplary brain sections and the visualization of the distribution of the two first components from a principal component analysis (PCA) of the image features. For the PCA, the same features were extracted as for the quantitative evaluation.

#### Evaluation with diagnosis classifier

Since the synthetic data was generated to improve the training of the diagnosis classifier, the final test for the synthetic data was to serve as training data for the classifier. The classifier was trained with a stratified 5-fold cross validation approach, using in each fold 60% of the data for training (61 SCZ and 55 HC images), and 20% of the real data for validation and test, respectively. Baseline classification accuracy was established training with real data only (Table 1). An additional baseline was constructed with an augmented data set of the same size as the real data training set. Here, we used the same augmentation techniques on the real images as for the DiffAugment step in the GAN evaluation: Random cropping of half the image size at most, translation of 1/8 maximum in each direction, and intensity randomization from 0.7 to 1-times the original value. Each image distortion was applied with a probability of one third.

**Table 1.**
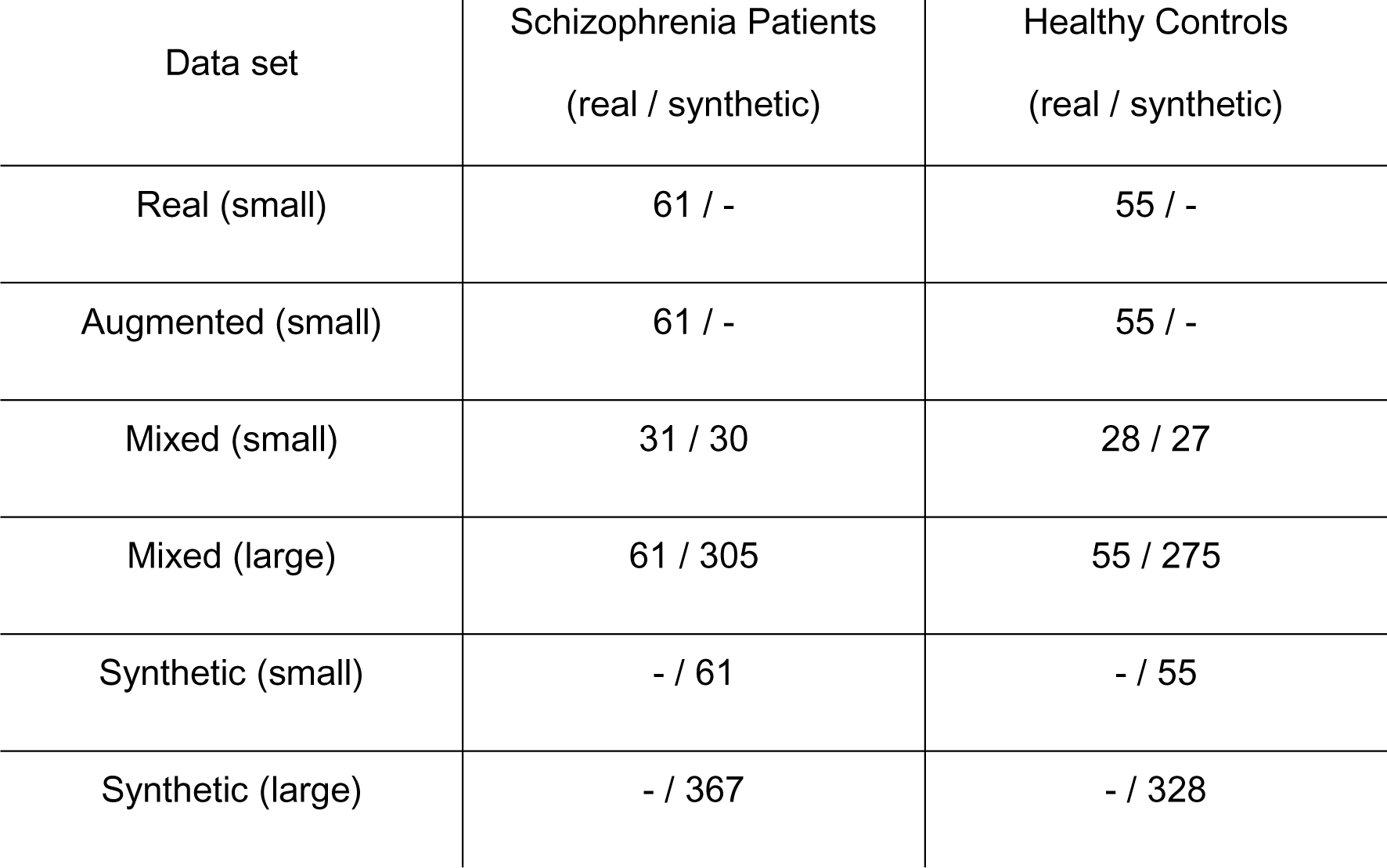
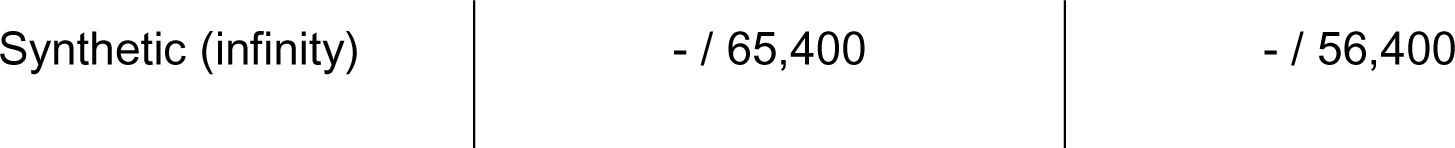
Training data sets for the diagnosis classifier.

In order to test the performance of training with synthetic data of the same size as the real training data, we constructed two training data sets containing synthetic images. For the mixed (small) data set, we replaced half of the real training data with synthetic data, for the synthetic (small) data, we replaced all real training data with synthetic images (Table 1).

Larger data sets are used for training to assess the effects of the data set size on the classifier’s performance. For the large mixed training data set, we boosted the real training data with five times the number of synthetic samples. This data set size is matched for the large synthetic training data set, containing synthetic images only (Table 1). Lastly, an “infinite” data set is trained, which is achieved by sampling a new random synthetic batch for each iteration. It reached its maximum performance after 4700 iterations, which equals a data set of 56,400 samples. Note that the validation and test data always consists of real data only, i.e. we test the robustness of the classifier to correctly classify real unknown MR images.

### Statistical analyses

For comparison of results produced by different generative models or for different data sets we use fixed-effects ANOVAs with between-subjects factors. Post-hoc tests are two-tailed unpaired *t*-tests with Bonferroni correction for all possible combinations of tests. Test results are reported as significant for α<.05. Reported values denote mean with distance to the upper and lower boundary of the 95% confidence interval.

## Results

### Basic architecture selection

All four quantitative evaluation metrics (Figure 2) perform sign. better without *DiffAugment* than with (all *F_1,72_*>35.21; *p*<.001) and demonstrate sign. differences between the architectures (all *F_3,72_*>317.03; *p*<.001). The average values of the evaluation metrics consistently show an advantage of the α-SN-GAN architecture over the three other models (Figure 2). Indeed, 10/11 out of the 12 post-hoc tests comparing this architecture to the other ones confirm a sign. better performance (all 11 tests *t_38_*>2.73; *p*<.010; sign. on α<.05 with/without correction for multiple comparisons).

**Figure 2.**
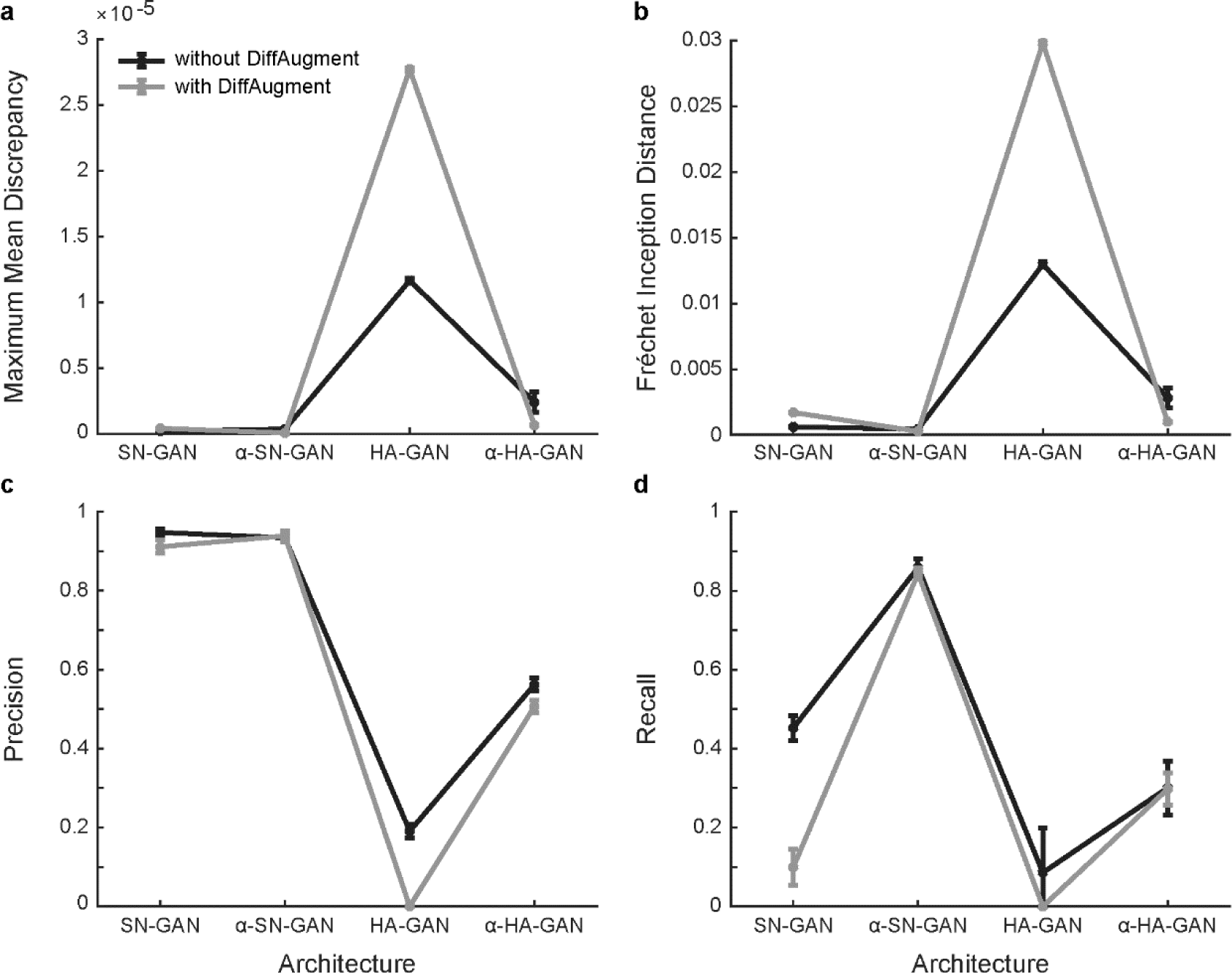
Quantitative evaluation metrics for the four basic architectures. Note that lower values indicate a better performance for MMD (a) and FID (b) while higher values indicate a better performance for precision (c) and recall (d). Error bars denote 95% confidence intervals. Abbreviations: SN (spectral normalization), GAN (generative adversarial network), α (with encoder), HA (hierarchical amortized).

Visually, the quality of the images synthesized by the SN-GAN and the α-SN-GAN is high (Figure 3). However, there are still limitations to fully resembling the real samples, especially regarding fine-grained structures. A clear difference between samples with and without DiffAugment is not present. The collapse of the hierarchical architectures is also visible here. Artefacts and deformations can clearly be seen in their samples. These appear stronger in the HA-GAN, which is in line with the findings of the quantitative evaluation. The illustrations of the PCA confirm these findings as well (Supplementary material Figure S7).

**Figure 3.**
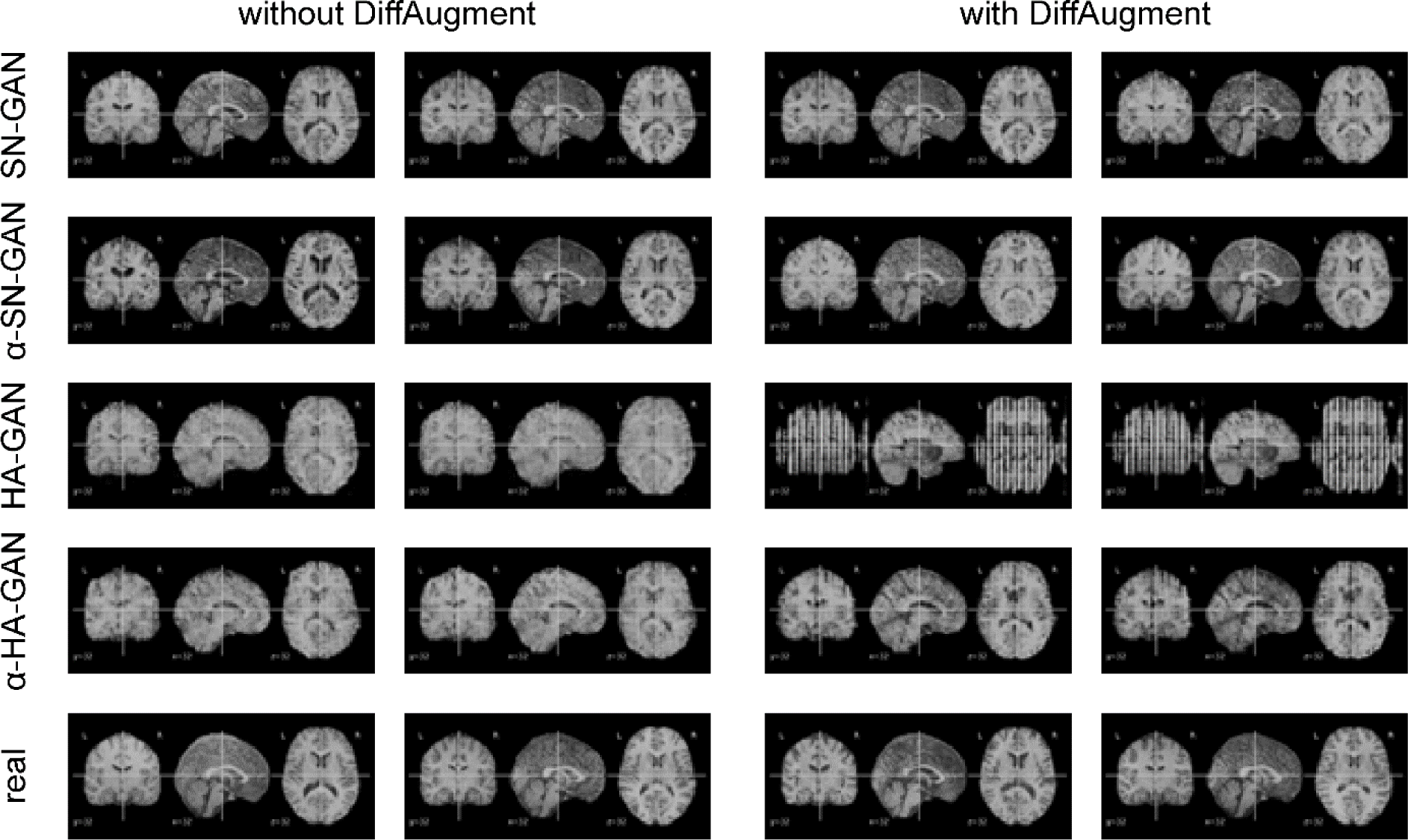
Exemplary synthetic images of the four architectures (rows) without and with DiffAugment applied (columns), two of each kind. Exemplary real images are presented in the bottom row for comparison. All sections show slice 32.

To summarize, regularization combined with incorporating an encoder (α-SN-GAN) yields synthetic images of highest fidelity and diversity shown with both qualitative and quantitative evaluation. The α-SN-GAN architecture without *DiffAugment* is therefore chosen as basis for the subsequent further architecture selection and data generation processes.

### Conditional architecture selection

All four quantitative evaluation metrics (Figure 4) demonstrate sign. differences between the conditional architectures *(F_2,54_*>10.06; *p*<.001) as well as the clinical groups (*F_1,54_*>11.40; *p*=.001). For the clinical groups, we do not find a clear advantage of one over the other: MMD and recall achieve a better performance for the synthetic patient images while FID and precision favour the synthetic HC images. For the conditional architectures, however, we find a consistent advantage of the architecture with the auxiliary classifier across all four evaluation metrics (Figure 4). Indeed, 6/7 out of the 8 post-hoc tests comparing the average values of this conditional approach to the other two confirm a sign. better performance (all 7 tests *t_38_*>2.97; *p*<.006; sign. on α<.05 with/without correction for multiple comparisons).

**Figure 4.**
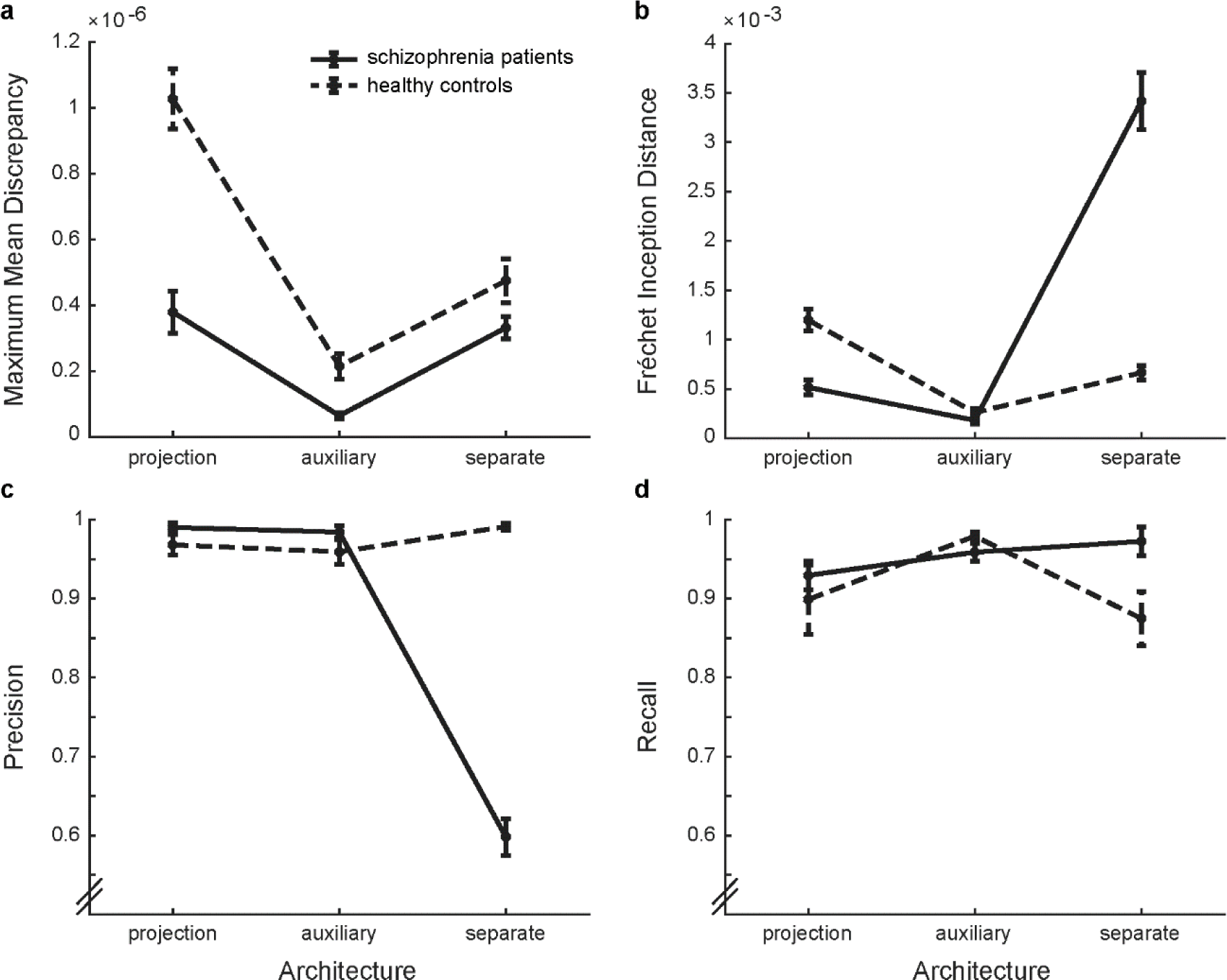
Quantitative evaluation metrics for the three conditional architectures. Note that lower values indicate a better performance for MMD (a) and FID (b) while higher values indicate a better performance for precision (c) and recall (d). Error bars denote 95% confidence intervals.

Visually, the quality of the generated images is high, rather independent of the conditional approach (Figure 5 and Supplementary material Figures S9 and S10). The illustrations of the PCA confirm the subtle differences found with the quantitative evaluation (Supplementary material Figure S8).

**Figure 5.**
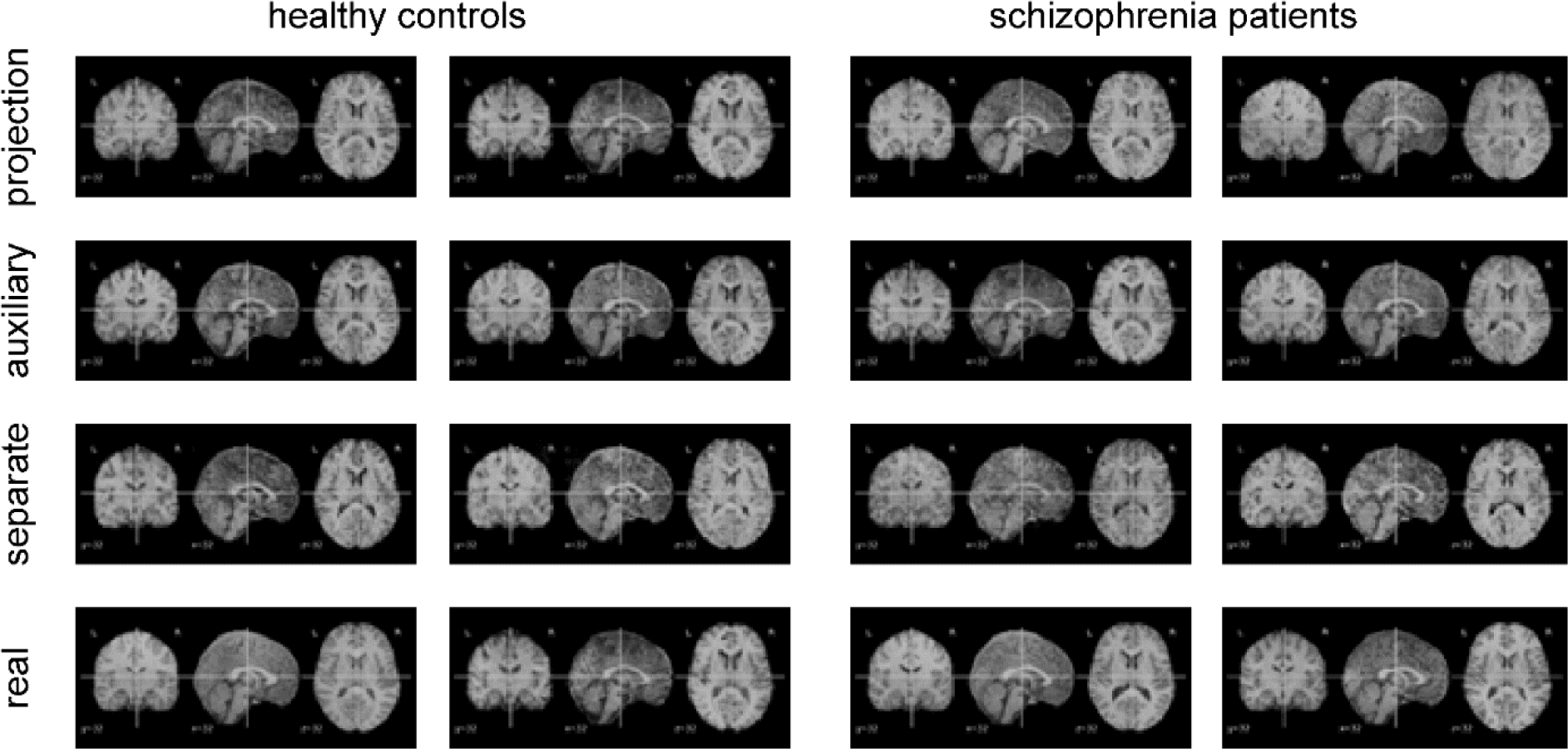
Exemplary synthetic images of the three differential conditional architectures based on the α-SN-GAN (rows) for the control and patient group (columns), two of each kind. Exemplary real images are presented in the bottom row for comparison. All sections show slice 32.

To summarize, synthesizing the two clinical groups with the auxiliary α-SN-GAN yields images of highest fidelity and diversity shown with both qualitative and quantitative evaluation. This architecture yields precision and recall values above 95% for both clinical groups (Figure 4c & d). The α-SN-GAN architecture with auxiliary classifier is therefore chosen as basis for the subsequent data generation processes.

### Diagnosis classification

Classification based on training with the real data achieves 60.6±7.6% diagnostic accuracy, a value well above chance level (*t_4_*=2.72; *p*=.026). However, this accuracy still leaves potential for improvement. Training the diagnosis classifier on the same amount of augmented, mixed, or synthetic data (Figure 6, small training data size) yields a comparable accuracy to training on the real data (*F_3,16_*=0.36; *p*=.786). Increasing the training data 6-fold yields sign. higher classification accuracy than training with the real data for both the mixed (*t_8_*=3.94; *p*=.002) and the synthetic (*t_8_*=4.06; *p*=.002) training data (Figure 6, large training data). Note that the classifiers are always tested on real data only. The strategy of the classifier is not biased towards the one or the other class when artificial data is added or used for training (cf. additional performance metrics in Supplementary Figure S11). Training with the 6-fold amount of synthetic data increased the diagnostic accuracy to 78.9±4.4%, which corresponds to an average increase of 18.3% with respect to the original classification accuracy of the classifier trained with real data only. Note that this accuracy cannot be increased further as demonstrated with the synthetic infinity training data set (Figure 6, diamond).

**Figure 6.**
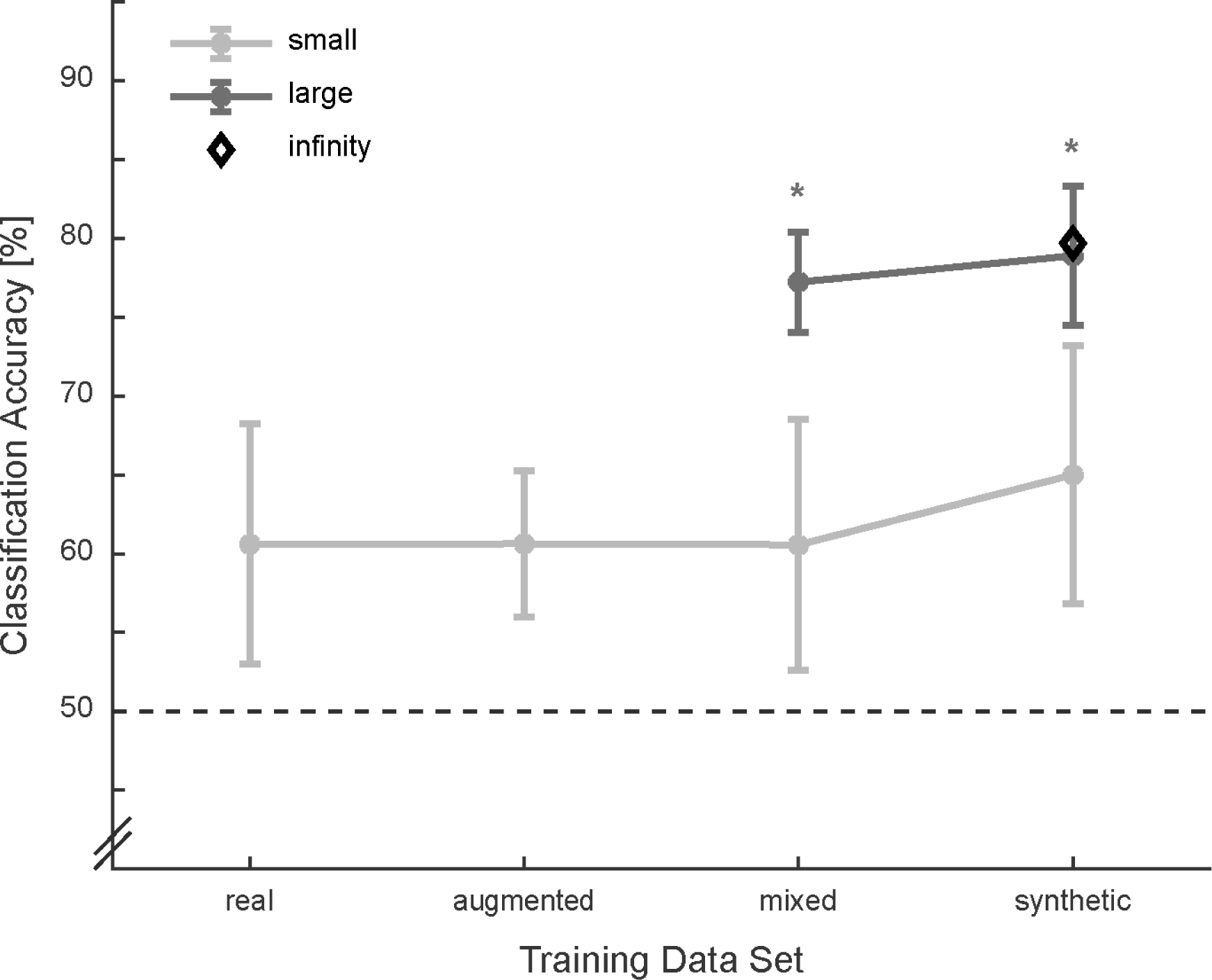
Classification accuracies for all training data sets. Note that the classifiers are always tested on real data only. Dotted line indicates chance level. Error bars denote 95% confidence intervals. * comparison to real data for training (Bonferroni corr. on α<.05).

## Discussion

This work demonstrates the synthesis of high-quality 3D brain sMRI data for two clinical groups from a very small data set. A diagnostic classifier separating real sMRI data from SCZ patients and HC can be trained with the synthetic data just as well as with the same amount of real data. Increasing the amount of synthetic training data 6-fold increases the performance of the diagnosis classifier by nearly 20%. This increase suggests that the synthetic data is capable of making the algorithm more robust for classifying the real data.

We demonstrate that regularized GANs produce high quality images even for small 3D sMRI data sets. Creating a high diversity of images instead of falling into mode collapse can be achieved by incorporating an encoder in the training process whereas applying DiffAugment to the GAN training did not improve image quality. Memory efficient processing with hierarchical architectures, however, was not successful in the case of the small data set used in this study. The study with the original implementation of the HA-GAN [14] used a larger data set of more than 3000 samples compared to the 193 images in this study. Considering that the HA-GAN and α-HA-GAN consist of five and eight individual neural networks, respectively, that all need to work in an equilibrium during the training, the low number of training samples might contribute to an unstable training process.

Comparing GAN metrics to related works is difficult due to multiple possible implementations of the MMD metric and additionally due to the use of a different feature extraction network. At least visually, however, the comparison can be made. The synthetic images in this study are on the same level as the ones of Kwon et al. [13] with the same image size but with a larger data set of around 500 images.

The first work demonstrating the feasibility of generating 3D sMRI data with a true 3D GAN was based on a subset of nearly thousand images [14]. Since the data included HC only, the usefulness of the data for clinical application could not be assessed. The first work conditioning the generated data for a clinical use case utilized a 3D architecture only for the discriminator but not for the generator, which constructed contingent 2D slices instead of volumes [17]. In the aforementioned study, the data quality was assessed thoroughly by statistical comparison of known disease biomarkers between the groups of generated data. A downstream diagnostic classifier, i.e. assessing the usefulness of the generated 3D data for training an algorithm that distinguishes between healthy and diseased persons, has not been tested yet. Indeed, differentiated MR images for SCZ patients and HC have not yet been synthesized either. The performance gain of the diagnosis classifier with the large synthetic data set is in the same range as found in a study using 2D MR images [48]. In that study, classification of Alzheimer’s patients and HC increased in accuracy from 63% to 83%. However, the data set consisted of slightly above 1000 samples compared to the 193 samples in this work. Admittedly, the diagnosis classifier’s performance of this study starts at the lower end of the spectrum but rises to the middle to upper end by augmenting with synthetic samples [8]. The increase is mainly contributed to the increased data set size combined with the high fidelity and diversity of the synthetic images. This is especially remarkable because classifier based on large data sets often do not reach a classification accuracy compared to small, specialized sets [10].

The systematic comparison of GAN architectures for basic training as well as for conditioning the data on the clinical group demonstrates that the architectural choices for the GAN are essential and the resulting data always needs to be evaluated carefully. Even though the α-SN-GAN architecture with auxiliary classifier generated the best synthetic data for the data set used in this study, this architecture may not be the best one for every data set and every purpose. We therefore suggest that always several architectural variants that address potential problems arising from the specific data set at hand are compared and thoroughly evaluated for the purpose the data is needed. Techniques that work for one application are not necessarily successful in another one despite their theoretical validity. Additionally, combinations of techniques may not be compatible.

This approach can also be adapted to bolster other imaging modalities such as functional MRI for training multimodal classifiers that have shown promise for SCZ diagnosis. Furthermore, the auxiliary classifier approach has the potential to reveal the underlying structural differences between two clinical groups and might therefore aid in the research for SCZ biomarkers. GANs are only one kind of generative DL architecture for image synthetisation. Other DL architectures such as transformers [49] have risen the past years and demonstrated exceptional capabilities in various domains not only language understanding and synthesis but also in medical image synthesis [50]. However, the problem of small medical data sets is even graver with these architectures and pre-training on data of other domains might have its limitations in medical application [51].

To conclude, generating synthetic (neuro)imaging data is a promising approach, especially for clinical use cases with inherently small data set sizes. With this work, we demonstrate the ability to train GANs even on a complex, small data set for a psychiatric disorder such as SCZ that lacks objective diagnostic tools. Importantly, the generated data enables a more robust training of a downstream diagnostic classifier.

## Data Availability

Data is available via the MCIC collection (https://www.nitrc.org/projects/mcic/).

## Acknowledgements

YH is supported by grant KK5207801BM0 from the Federal Ministry for Economic Affairs and Climate Action (BMWK) on the basis of a decision by the German Bundestag. We thank F. Greiner for his help in data pre-processing.

## Author contribution statement

**Sebastian King**: Methodology, Software, Verification, Formal analysis, Investigation, Writing – Original Draft, Visualization. **Yasmin Hollenbenders**: Methodology, Software, Investigation, Data Curation, Writing – Review & Editing, Supervision. **Alexandra Reichenbach**: Conceptualization, Formal analysis, Resources, Writing – Review & Editing, Visualization, Supervision, Funding acquisition.

## Additional information

The authors declare no competing interests.

## Summary

Schizophrenia and other psychiatric disorders can greatly benefit from objective decision support in diagnosis and therapy. Machine learning approaches based on neuroimaging, e.g. magnetic resonance imaging (MRI), have the potential to serve this purpose. However, the medical data sets these algorithms can be trained on are often rather small, leading to overfit, and the resulting models can therewith not be transferred into a clinical setting. The generation of synthetic images from real data with generative adversarial networks (GAN) is a promising approach to overcome this shortcoming. Due to the small data set size and the size and complexity of medical images, those algorithms are challenged on several levels:

1) small data sets can lead to discriminator overfit, causing the vanishing gradients problem 2) most algorithms for image processing were first, or only, implemented and optimized for 2D data; 3) complex, i.e. 3D, images tend to converge to a very small distribution of generated images causing mode collapse; 4) 3D images and operations need exponentially more memory.

We addressed these challenges with the development and comparison of four GAN architectures and compared them systematically based on a data set of 193 MR images of schizophrenia patients and healthy controls. Spectral normalization regularization (SN-GAN) counteracts the vanishing gradients problem and is applied for all architectures. Additionally incorporating an encoder (α-SN-GAN) helps to alleviate mode collapse. To reduce the computational cost of the training, a hierarchical approach is adapted (HA-GAN), which is also combined with the α-SN-GAN to join their advantages (α-HA-GAN). Vanishing gradient and mode collapse are additionally addressed by applying data augmentation during training to all four architectures. The best architecture (α-SN-GAN without augmentation) is selected for further processing based on qualitative and quantitative evaluation of the generated images.

Subsequently, three conditioning approaches are employed for creating images of the two clinical groups: one classifier per class, an auxiliary classifier, and a projection discriminator. The winner architecture (α-SN-GAN with auxiliary classifier) is then used to generate different sets of training data with different ratios of real and synthetic data and different set sizes.

Finally, a diagnosis classifier is trained on these data sets to separate patients from controls in a test data set consisting of real data only. The synthetic images increase the accuracy of the diagnostic classifier from a baseline accuracy of around 61% to 79%.

This study demonstrates for the first time the end-to-end generation of high-quality synthetic data from a very small 3D sMRI data set with a true 3D GAN and downstream data-driven diagnosis classification. Additionally, this is also the first time to produce 3D MR images for SCZ classification.

## Supplementary material

### Architecture diagnosis classifier

**Figure S1.**
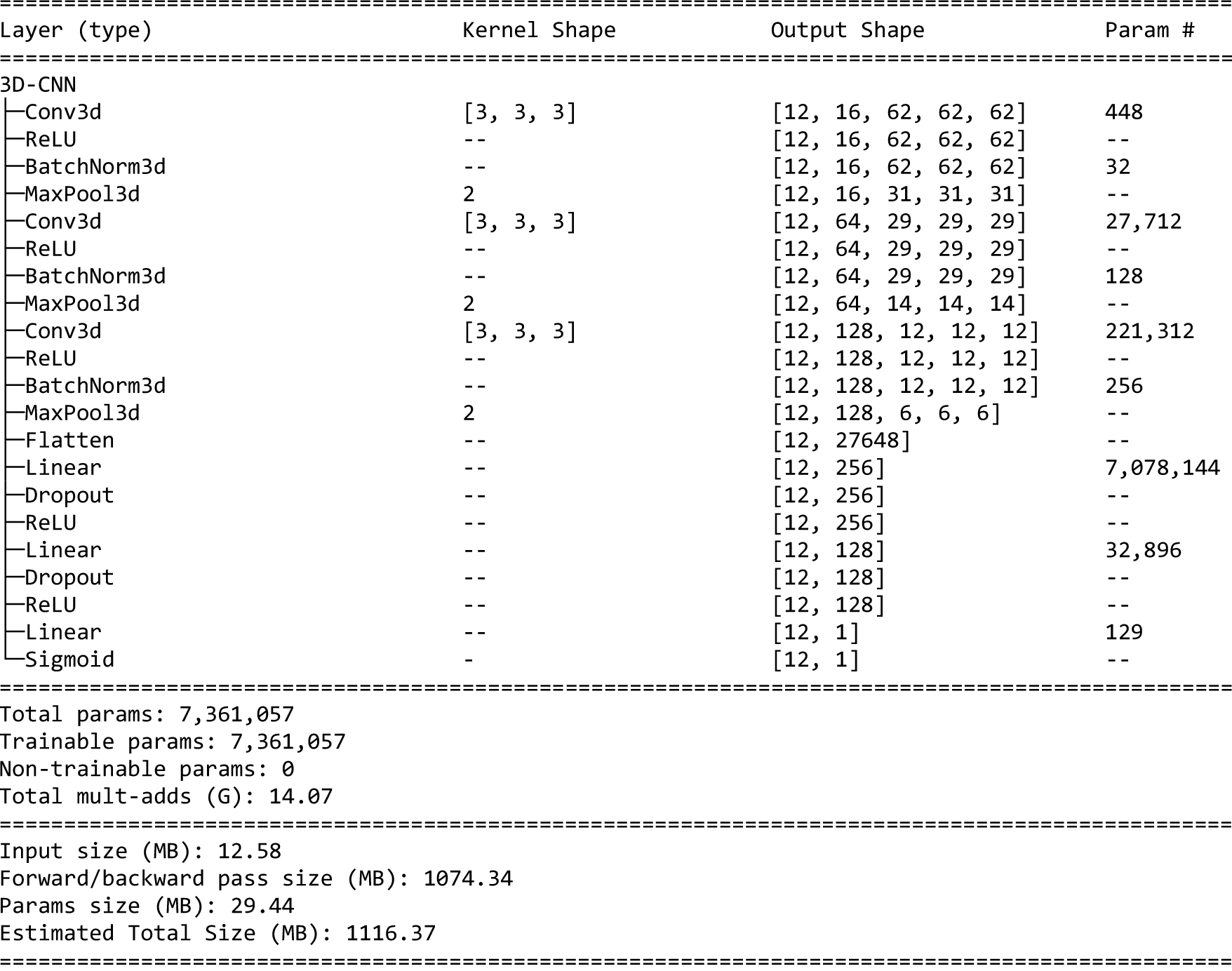
Architecture of the 3D-CNN used for the diagnosis classifier.

### GAN architectures

**Figure S2.**
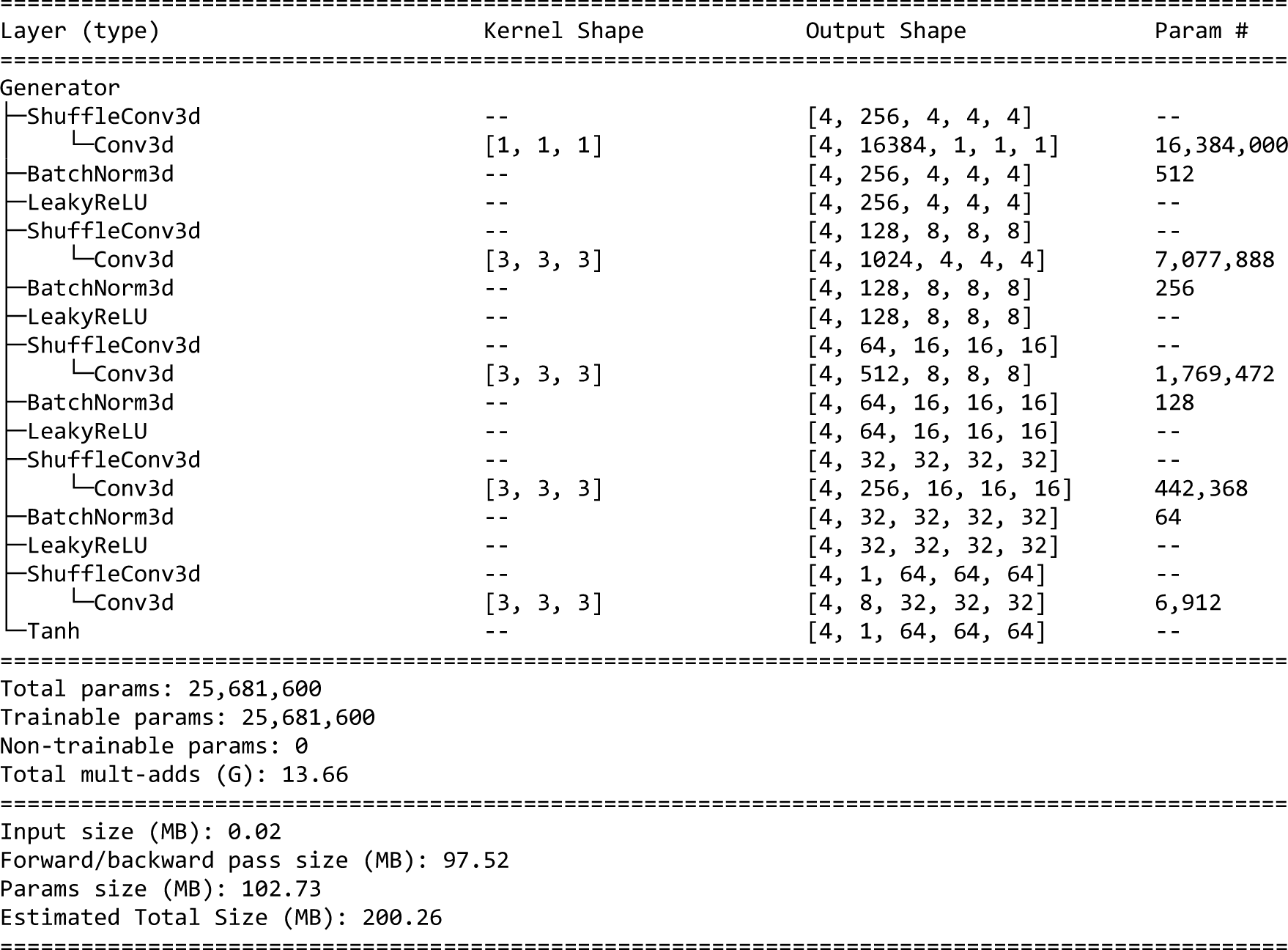
Generator architecture of the SN-GAN and α-SN-GAN. For the hierarchical approaches, the generator is split after the third convolution.

**Figure S3.**
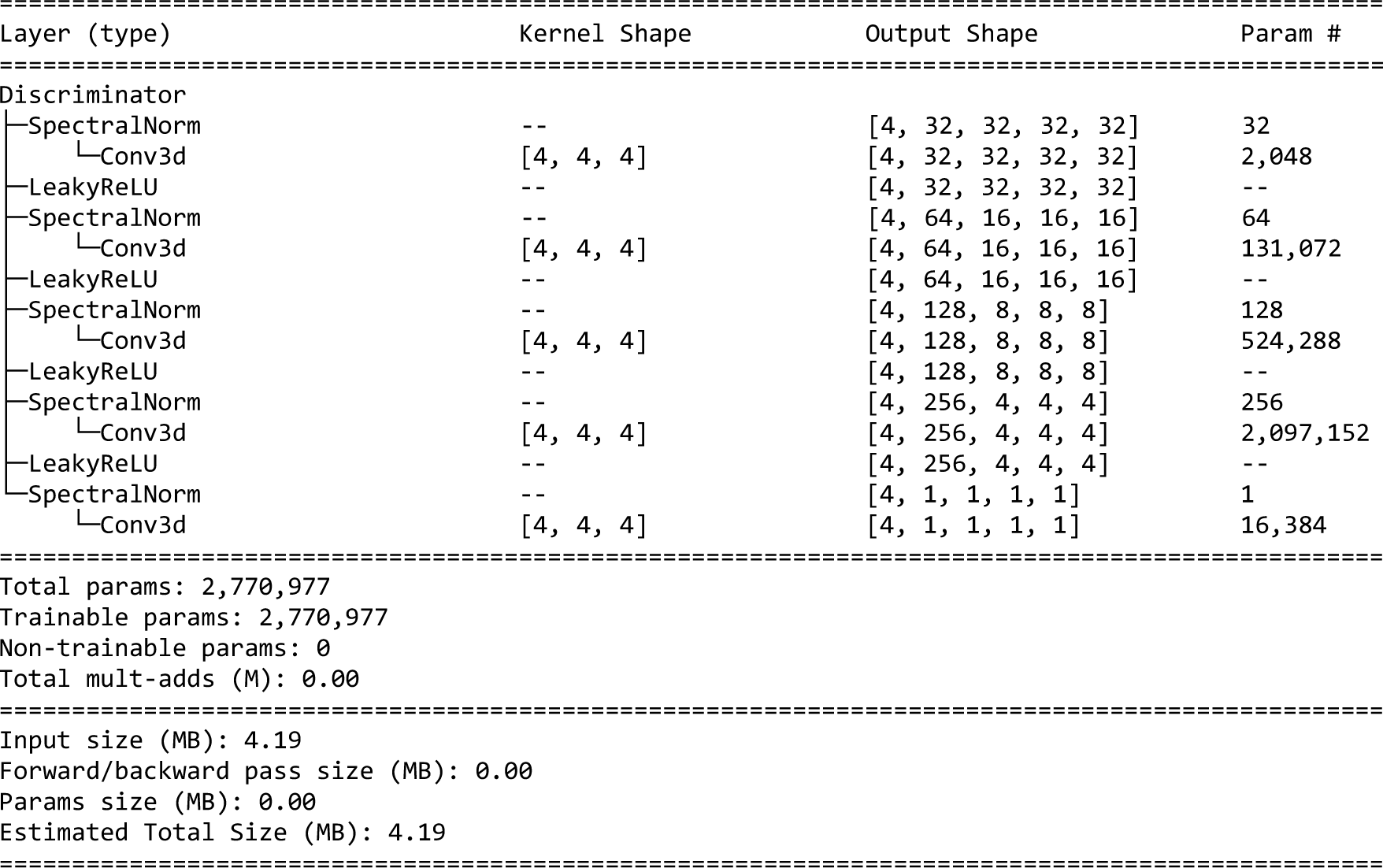
Discriminator architecture of the SN-GAN, the α-SN-GAN, and the auxiliary classifier. For the hierarchical approaches, the discriminator is split after the third convolution.

**Figure S4.**
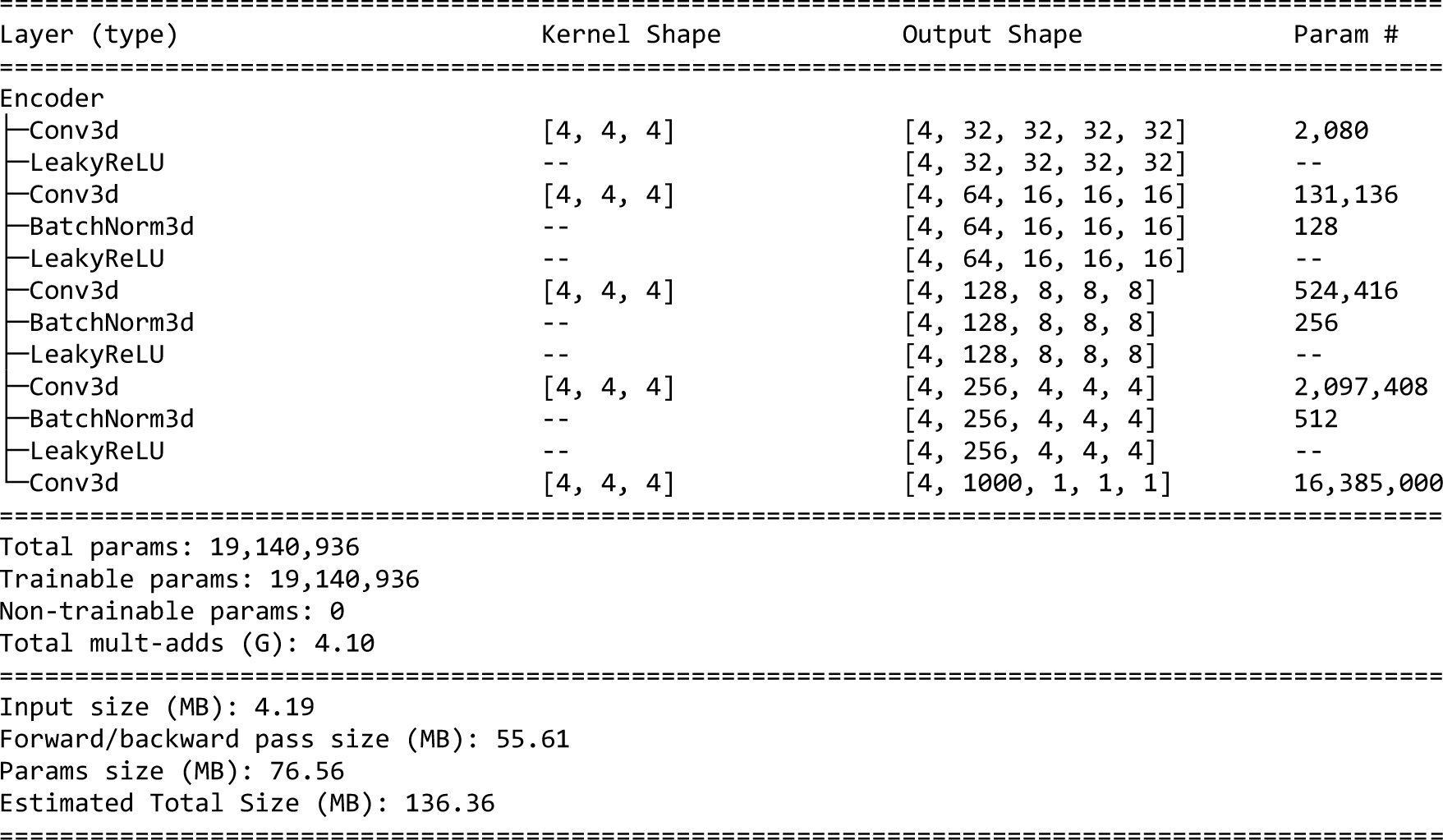
Encoder architecture of the α-SN-GAN. For the hierarchical approaches, the encoder is split after the second convolution layer to match its output size with the input of the high-resolution generator.

**Figure S5.**
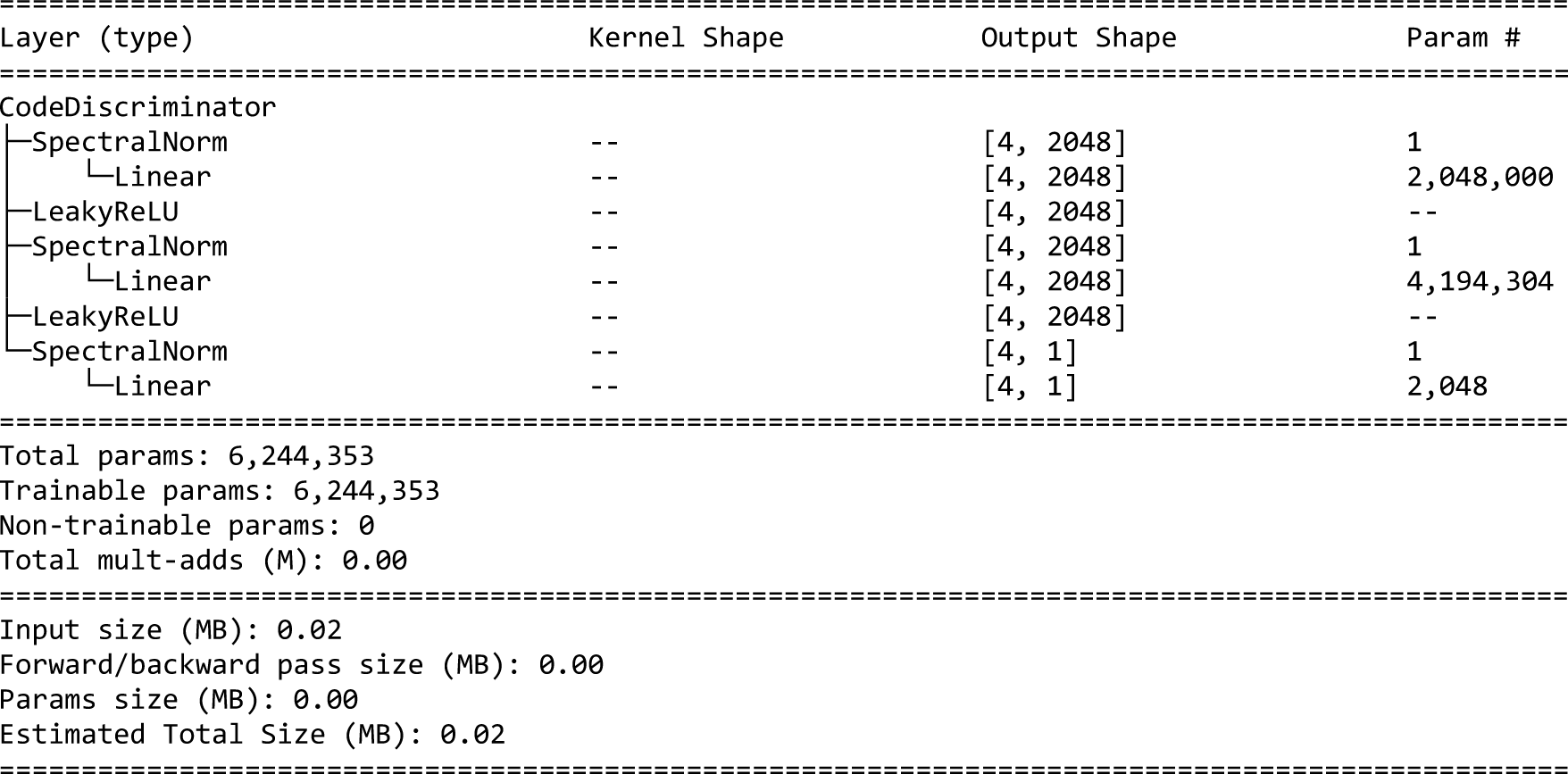
Code discriminator of the α-SN-GAN and the α-HA-GAN.

### α-HA-GAN

Eight neural networks are required for training the α-HA-GAN (Figure S6). The shared generator is defined as G^A^ whereas the high- and low-resolution generators are defined as G^H^ and G^L^. Discriminators and encoders are defined as D^H,^ D^L^, E^H^ and E^L^ respectively. Finally, the code discriminator network is described as CD. Randomly sampled latent space are defined as z_r_, real images as X, low-resolution ones as X^L^, and sub-volumes as X^H^. Encoder outputs are labelled X_e_ for E^H^ and z_e_ for E^L^. Both G^H^ and D^H^ receive the sub-volumes position c as additional input. Discriminators are combined by averaging their outputs so that D(∗, c) = {(*D*}^*H*^(∗, c) + *D*^*L*^(∗)) / 2. During the training, the generator models G^A^, G^L,^ and G^H^ are also combined to form a single network G, which outputs both a synthetic small image 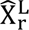 and a synthetic sub-volume 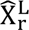 for z_r_. Reconstructed image outputs when using z_e_ are labelled 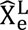 and 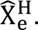 Since E^L^ and G can be treated as one network in the training their loss functions are combined (L_EG_), which results in four loss functions (Equations 1 - 4).

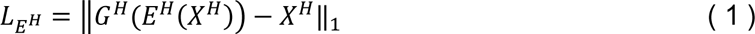

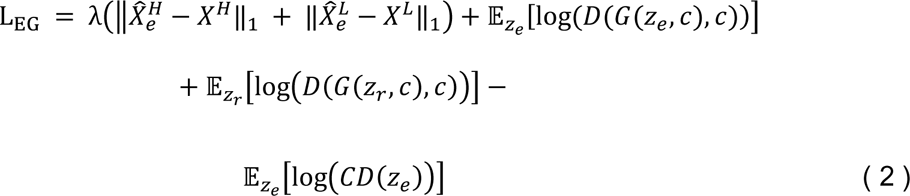

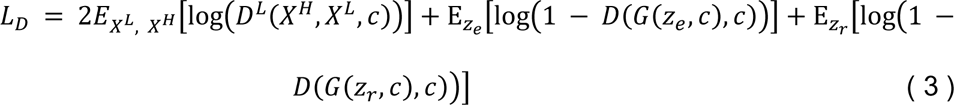

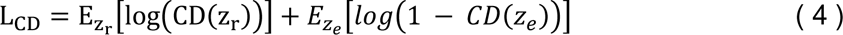

**Figure S6.**
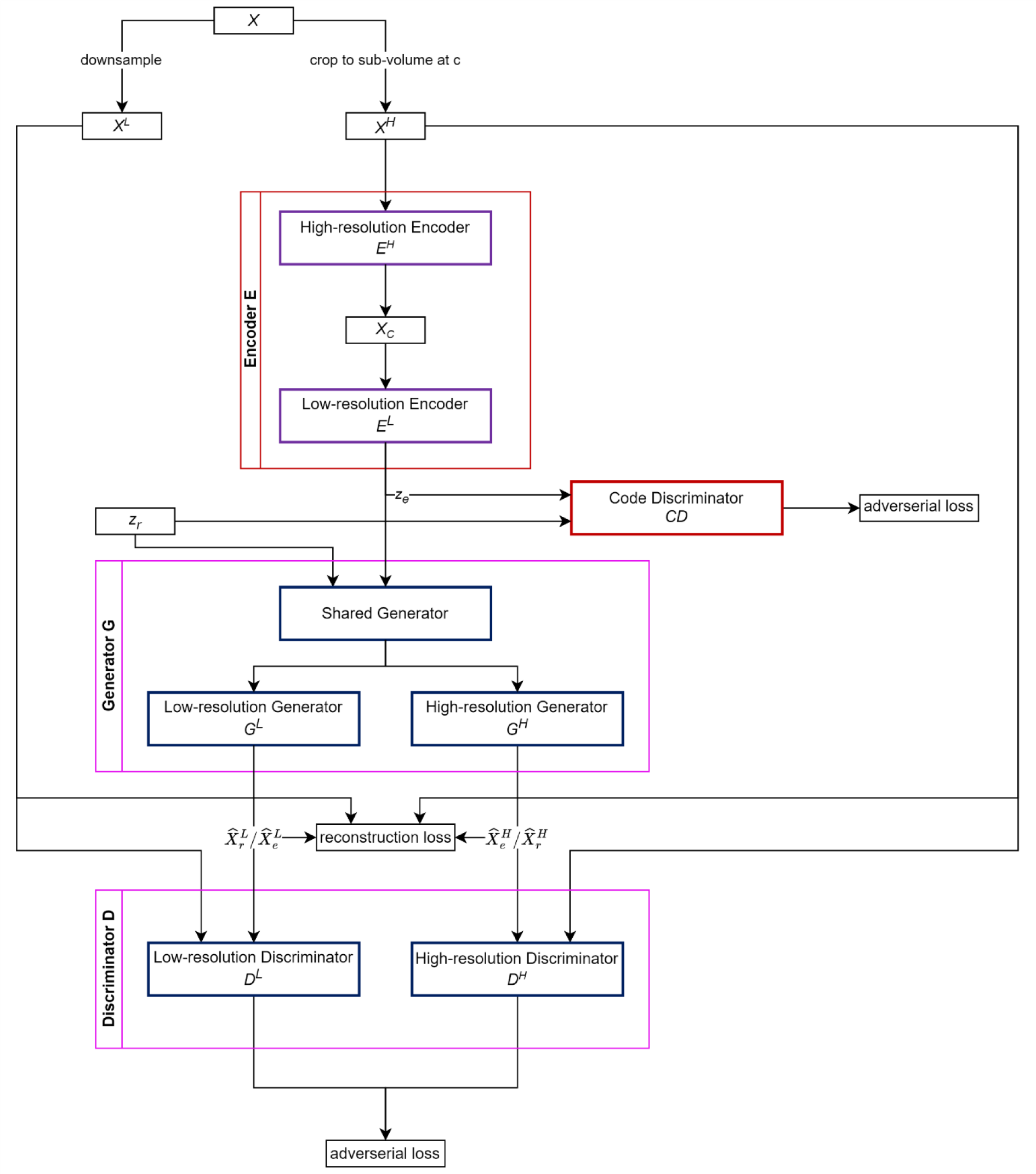
α-HA-GAN architecture including the submodels of the combined discriminator and generator as well as inputs and outputs during training mode. Interestingly, all previously introduced architectures are included in the α-HA-GANs architecture as well (colour code according to Figure 1): Removing the Encoder E and the Code Discriminator CD produces the HA-GAN. Looking only at the combined networks E, CD, G, and D without the high-and low-resolution path, the α-SN-GAN architecture is formed. Removing E and CD from this subset results in the SN-GAN architecture.

**Figure S7.**
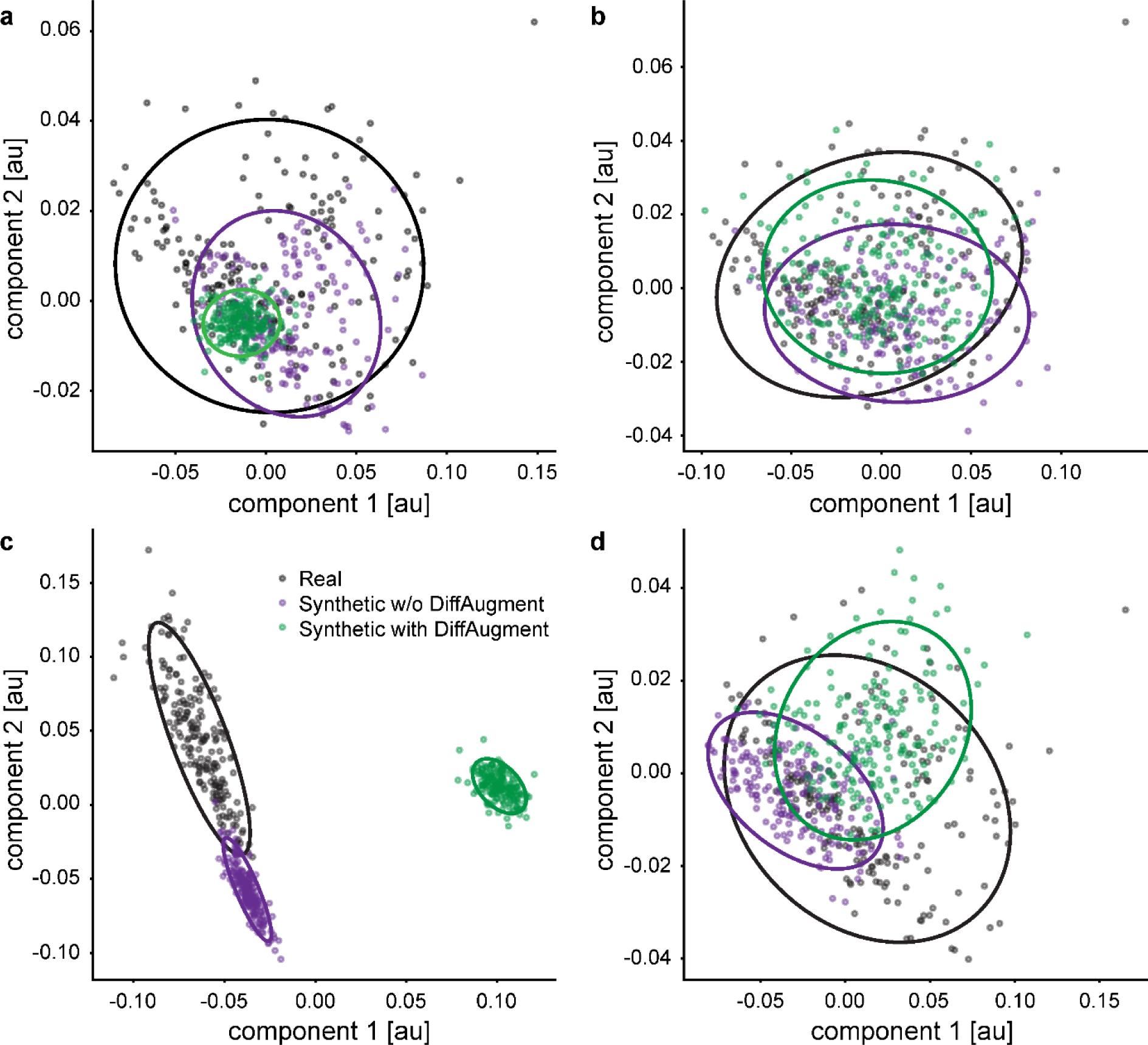
PCA results with the two first components for the a) SN-GAN, b) α-SN-GAN, c) HA-GAN, and d) α-HA-GAN. The analyses are based on all 193 real samples and an equal number of synthetic samples.

**Figure S8.**
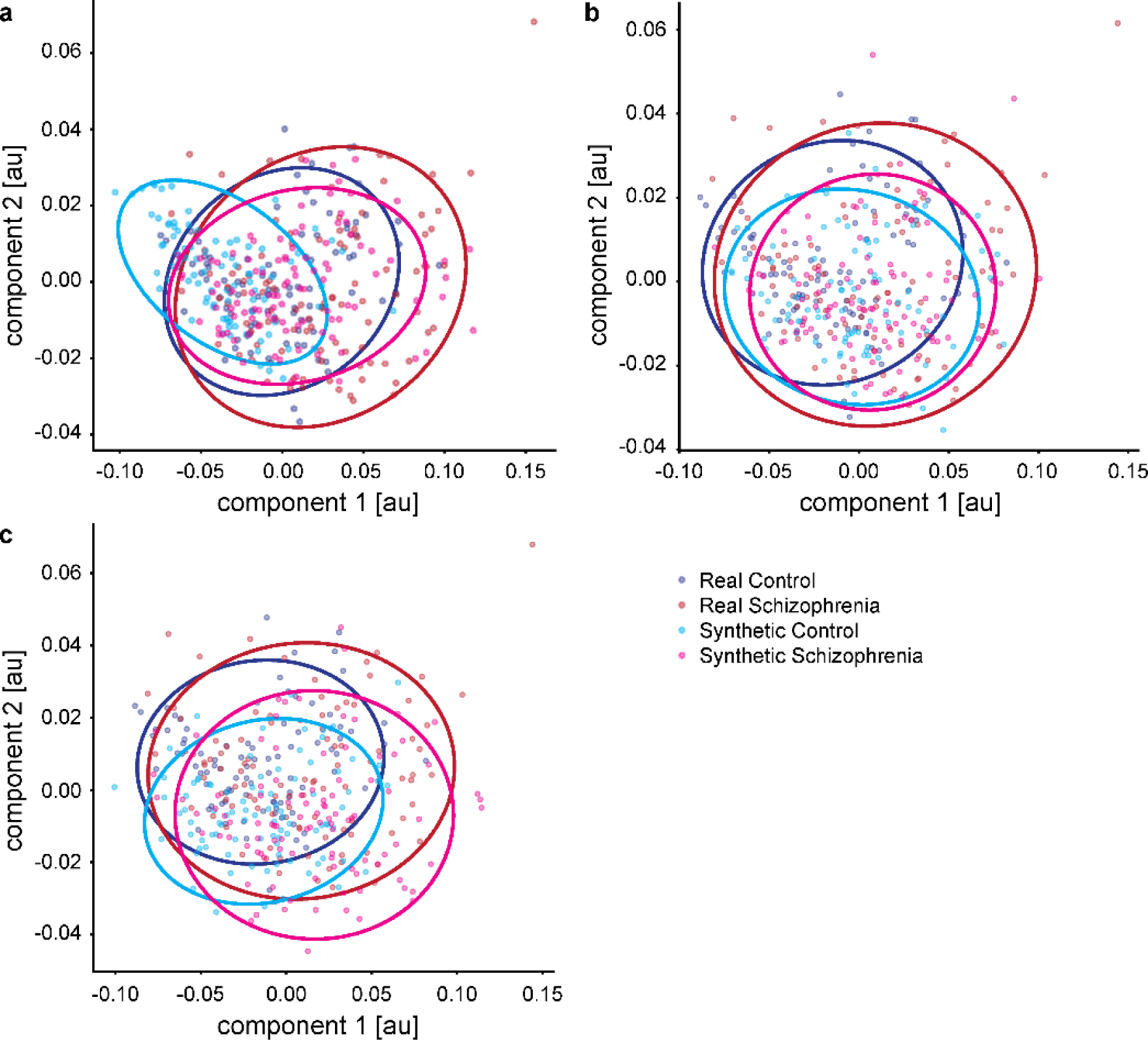
PCA results with the two first components for the a) projection discriminator, b) auxiliary classifier, and c) separate data synthesis. The analyses are based on all 193 real samples (91 healthy controls and 102 schizophrenia patients) and an equal number of synthetic samples.

**Figure S9.**
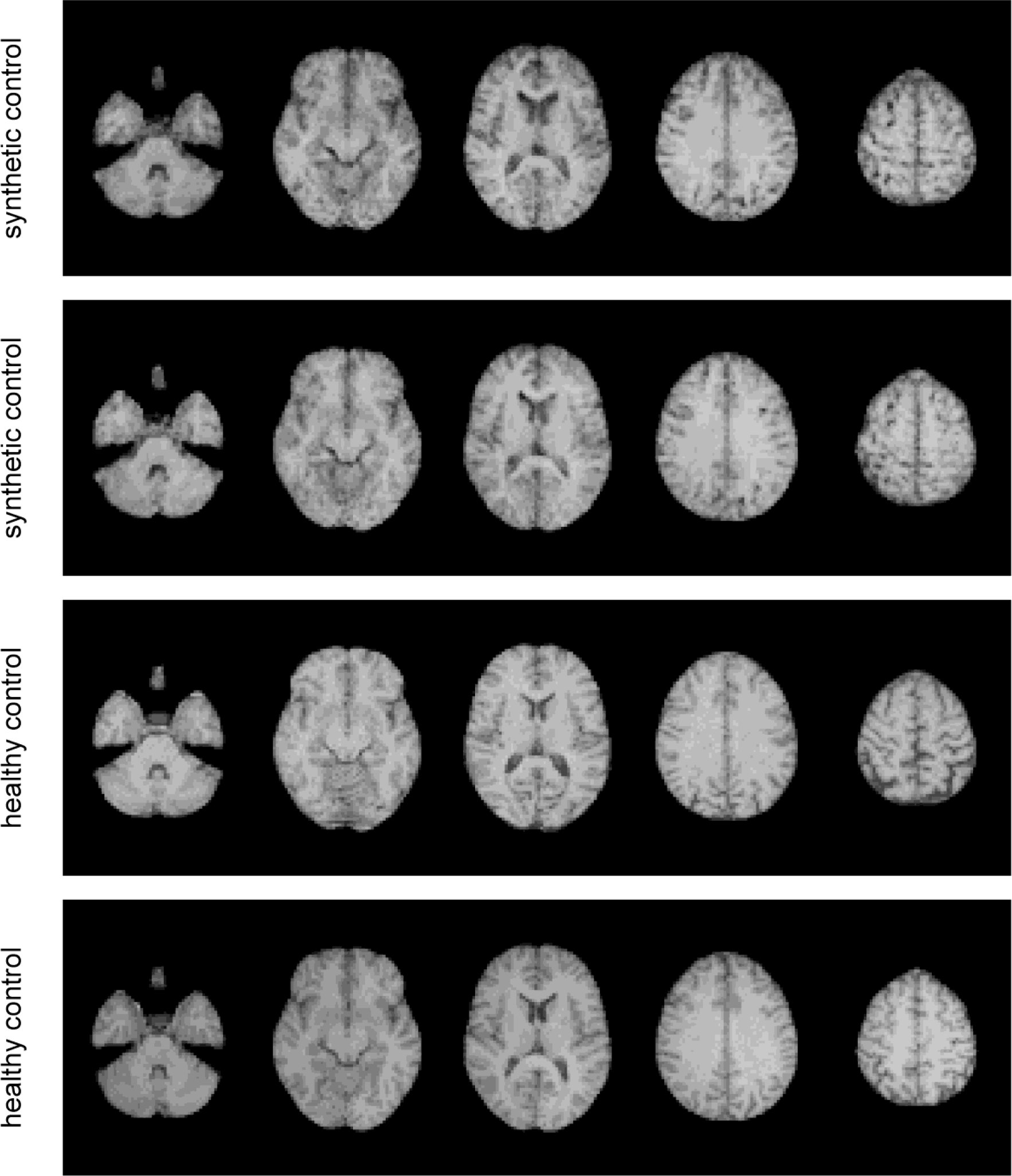
Exemplary synthetic (top rows) and real (bottom rows) images from healthy control subjects, or the respectively labelled group of synthetic data. The synthetic images are generated with the α-SN-GAN without DiffAugment and with the auxiliary classifier for generation of group-specific images. i.e. with the winning architecture used for synthetizing data for the diagnosis classifier. The horizontal sections show slices 16, 24, 32, 40, and 48.

**Figure S10.**
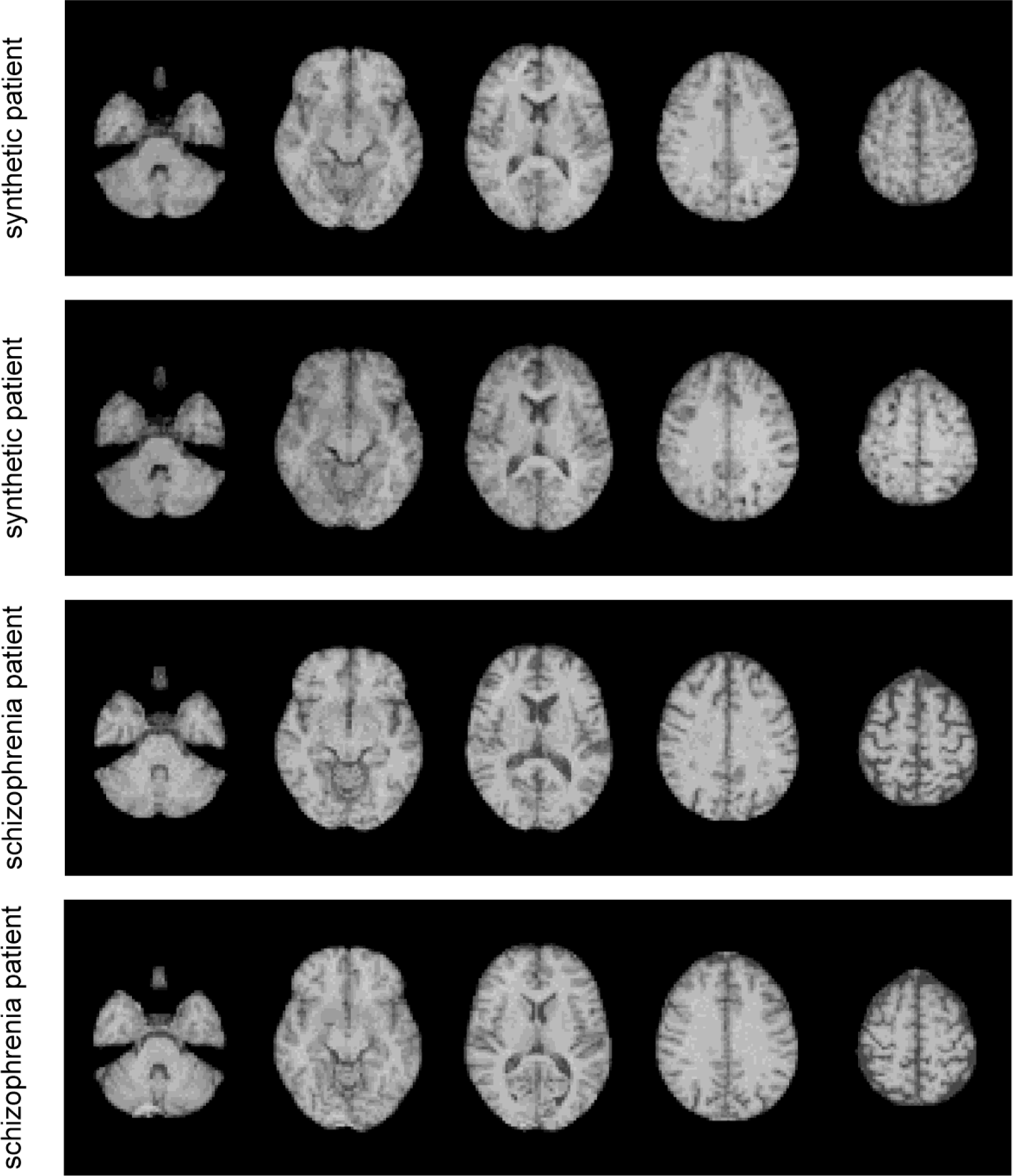
Exemplary synthetic (top rows) and real (bottom rows) images from schizophrenia patients, or the respectively labelled group of synthetic data. The synthetic images are generated with the α-SN-GAN without DiffAugment and with the auxiliary classifier for generation of group-specific images. i.e. with the winning architecture used for synthetizing data for the diagnosis classifier. The horizontal sections show slices 16, 24, 32, 40, and 48.

**Figure S11.**
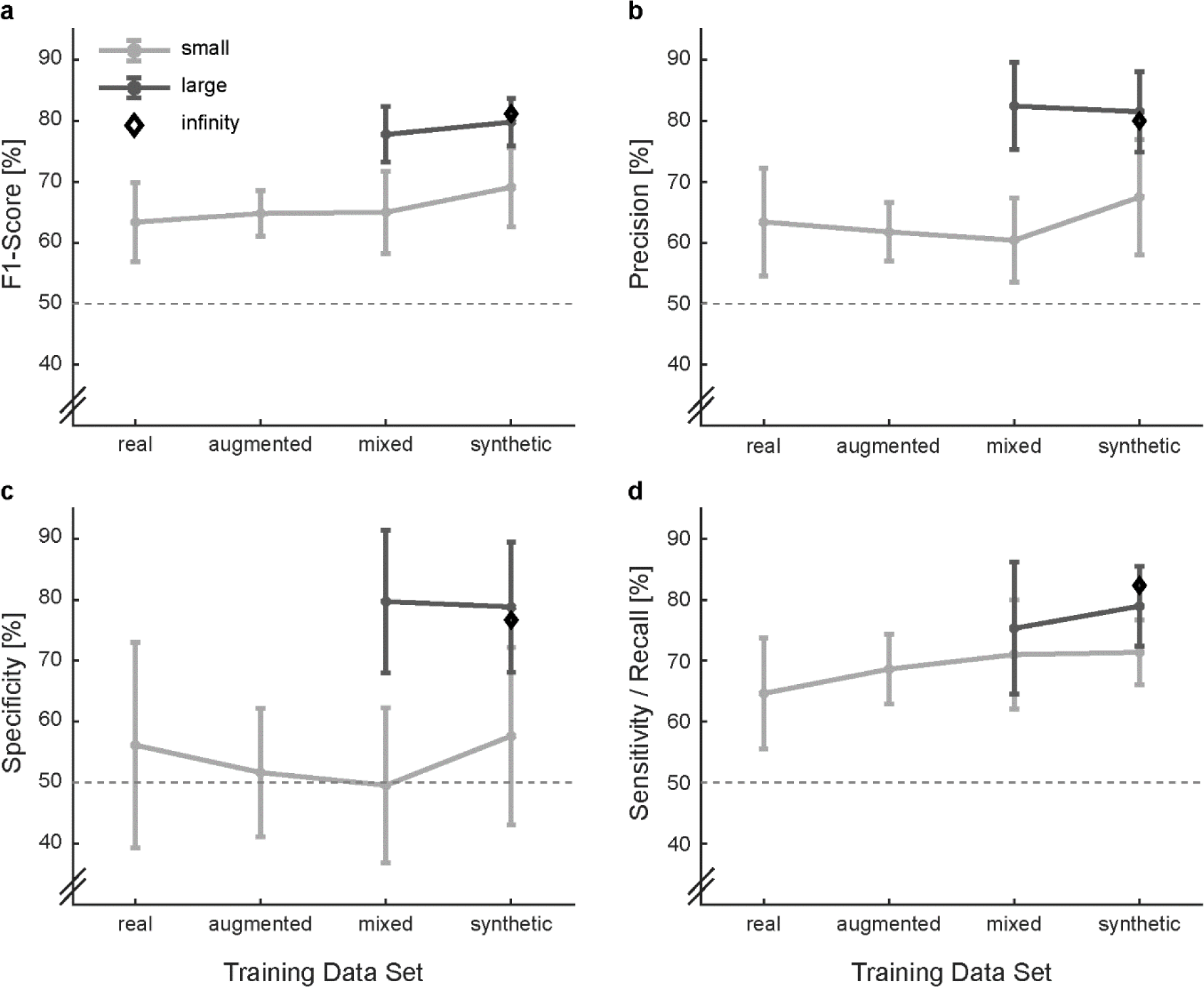
Additional classification evaluation metrics for all training data sets. Note that the classifiers are always tested on real data only. Dotted lines indicate chance level. Error bars denote 95% confidence intervals.

